# Hand–Washing Hygiene Knowledge, Attitude and Practice among University Students in the Post-COVID-19 Era: A Case of KCMC University, Moshi, Tanzania

**DOI:** 10.64898/2026.10.09.26365114

**Authors:** Emmanuel T. Issangya, Victor V. Mosha, Dhahiri Mnzava, Rhodes Mwageni, Debora C. Kajeguka

## Abstract

**Background:** Hand hygiene is a key public health intervention for preventing infectious diseases, including respiratory and gastrointestinal infections. Although the COVID-19 pandemic improved awareness and compliance, sustained adherence remains a challenge, with evidence showing a decline in practice after outbreaks, particularly among young adults in high-contact settings such as universities. Therefore, this study aimed to assess the knowledge, attitudes, and practices of handwashing hygiene among university students at KCMC University, Moshi, Tanzania, in the post-COVID-19 era.

**Methods:** A cross-sectional study was conducted among 380 students at KCMC University. Data were collected using a pretested structured questionnaire, which captured information on socio-demographic characteristics, knowledge, attitudes, and practices regarding handwashing hygiene. Data were analyzed using descriptive and inferential statistics. Associations between variables were assessed using logistic regression and multivariable analysis, and a *p*-value of <0.05 was considered statistically significant.

**Results:** A total of 380 students participated in the study, with a mean age of 23.5 ± 3.5 years. More than half 209 (55.0%) were aged 18–23 years, while 203 (53.4%) were female. Most participants had access to hand hygiene facilities 338 (88.9%). 338 (88.9%) participants had good knowledge of hand hygiene but attitudes were mostly negative 193 (50.8%), and practices were suboptimal, with 201 (52.9%) showing poor hand hygiene practices. Good hand hygiene knowledge was associated with access to hand hygiene facilities (AOR=2.84, 95% CI: 1.25– 6.42, *p*=0.012) and formal training on hand hygiene (AOR=3.48, 95% CI: 1.24–9.76, *p*=0.018). Positive attitudes were associated with being in the fourth year or above (AOR=2.12, 95% CI: 1.16–3.89, *p*=0.015). Good hand hygiene practices were associated with access to hand hygiene facilities (AOR=2.67, 95% CI: 1.18–6.01*, p*=0.018) and formal training on hand hygiene (AOR=2.98, 95% CI: 1.19–7.48, *p*=0.020).

**Conclusion:** Although knowledge and awareness of handwashing hygiene were generally high among university students, inconsistencies between knowledge and actual practices were observed in the post-COVID-19 era. Strengthening health education programs and ensuring continuous access to handwashing facilities may improve adherence to proper hand hygiene practices among university students.

## Background

Hand Washing hygiene is a process of cleansing hands with soap and running water and drying to remove dirt, grease and pathogenic microorganisms such as bacteria and viruses as intervention of the spread of infectious diseases and infections transmitted through respiratory droplets, direct contact, and the fecal–oral route [1,2]. Hand Washing hygiene is the most effective and cost-efficient intervention for preventing infectious diseases [3,4].

Hand hygiene gained unprecedented attention during the Coronavirus Disease 2019 (COVID- 19) pandemic, a severe Acute Respiratory Syndrome Coronavirus 2 (SARS-CoV-2). The spread of COVID-19worldwide, prompted public health to recommend frequent hand washing, use of alcohol-based hand sanitizers, physical distancing, and mask use as key preventive interventions [1,5]. Hence increased knowledge, attitude and practices across different institutions, workplaces, healthcare facilities, and public spaces [6]. Universities reported varying levels of knowledge, attitudes, and practices regarding hand hygiene [7]. The majority of students demonstrated adequate knowledge about the importance of hand washing. Promotion campaigns were posted and hand washing facilities were installed to reduce COVID-19 pandemic in University settings [8,9]. In Tanzania, health education campaigns and the provision of hand washing facilities were improved during COVID-19 [10].

However, knowledge demonstrated by students does not always translate into appropriate practice [7,11,12]. This is because of poor hand hygiene practices include inadequate access to hand washing facilities, limited availability of soap and water, lack of institutional support, behavioral complacency, and low perception of susceptibility to infectious diseases [9,13].

It is evident that hand hygiene knowledge, attitude and practices increased during the COVID- 19 pandemic, but adherence to recommendations has declined in the post-pandemic period [9,14]. In LMICs there are gaps between knowledge and actual practices despite increased awareness campaigns [15–17]. Studies by WHO (2021a) WHO (2023), CDC (2024), Freeman *et al.* (2014), Okesanya, Eshun, Ukoaka (2024) affirm that inadequate hand hygiene remains a major public health challenge in which individuals often revert to pre-pandemic hygiene behaviors once the perceived threat of infection decreases. The practices contribute to the spread of infectious diseases and infections including diarrheal diseases, acute respiratory infections, influenza, cholera, typhoid fever, and gastrointestinal infections in Low- and Middle-Income Countries (LMICs) [18]. Furthermore, there is decimal evidence on the increased sustainability of hand washing knowledge (awareness), attitude (behavioral changes) and practices among university students observed during the COVID-19 pandemic have been maintained in the post-pandemic [19]. Understanding hand washing hygiene levels of knowledge, attitudes, and practices among university students is essential for identifying existing gaps and informing interventions in higher education. Therefore, this study aimed to assess the knowledge, attitudes, and practices of hand-washing hygiene among university students in the post-COVID-19 era.

## Methods and Materials

### Study Design and Study Period

This study employed a cross-sectional study design conducted among undergraduate students at KCMC University, Moshi, Tanzania. The study was conducted during the academic year from August 2025 to June 2026 and data were collected within the defined study period to capture current post-COVID-19 hygiene-related behaviors among university students.

### Study Area

The study was conducted at KCMC University, located in Moshi Municipality, Kilimanjaro Region, northern Tanzania. Geographically, University lies at approximately 3.35° South latitude and 37.33° East longitude, at the foothills of Mount Kilimanjaro. The area experiences a tropical climate with relatively moderate temperatures throughout the year. KCMC University is a higher learning institution situated within the Kilimanjaro Christian Medical Centre (KCMC) complex, which serves as a major referral and teaching hospital in northern Tanzania. The university offers undergraduate and postgraduate training in health and allied sciences and attracts students from different regions of Tanzania, creating a diverse academic population. The institution is characterized by a mix of on-campus academic buildings, student hostels, and nearby residential areas, where students engage in both academic and daily social activities. This setting provides an appropriate environment for assessing knowledge, attitudes, and practices of handwashing hygiene among university students in the post-COVID-19 era, particularly due to the shared facilities and high level of interpersonal interactions.

### Study Population

The study population comprised undergraduate students enrolled at KCMC University, Moshi, Tanzania, during the study period. These students represented various academic years and programs within the university. The study targeted students who were available at the time of data collection and who met the eligibility criteria, including provision of written informed consent.

### Eligibility Criteria

The study included undergraduate students enrolled at KCMC University, Moshi, Tanzania, during the study period. Eligible participants were those who were present at the time of data collection and who provided written informed consent to participate in the study. Students who declined to participate or who did not complete the questionnaire were excluded from the analysis. In addition, individuals who were not officially registered as students of KCMC University during the study period were also excluded.

### Sampling and sample size determination

A simple random sampling technique was used to select eligible participants from the list of registered students obtained from the university records to ensure equal probability of selection. The sample size was determined using the formula for a single population proportion, assuming a 95% confidence level, 5% margin of error, and an estimated proportion of 50% to maximize sample size due to the absence of prior similar studies among university students in the study setting. After adjusting for a 10% non-response rate, the final minimum required sample size was calculated accordingly. Eligible participants were proportionately recruited until the required sample size was achieved.

### Survey Questionnaire and Participants’ Recruitment

The survey questionnaire was developed based on an extensive review of relevant literature[15,20]. It was initially prepared in English, translated into Kiswahili, and then uploaded onto the KoboToolbox platform (https://www.kobotoolbox.org/) for electronic data collection. Participants were recruited using a structured approach to ensure eligibility and representation of university students at KCMC University, Moshi, Tanzania. The purpose of the study was clearly explained to all eligible participants, and written informed consent was obtained prior to participation. The questionnaire was organized into four main sections. The first section captured socio-demographic characteristics of respondents, including age, gender, level of education, marital status, and other relevant background information.

The second section assessed knowledge of handwashing hygiene, including awareness of handwashing as a key preventive measure against infectious diseases, critical moments for hand hygiene (such as before eating, after using the toilet, and after contact with contaminated surfaces), and understanding of disease transmission through poor hand hygiene practices. The third section evaluated attitudes towards handwashing hygiene, focusing on participants’ perceptions of the importance of handwashing, perceived susceptibility to infections due to poor hygiene, perceived effectiveness of handwashing in preventing disease transmission, and willingness to maintain proper hand hygiene practices in daily life. The fourth section assessed handwashing practices, including frequency of handwashing, use of soap and clean water, adherence to proper handwashing techniques, and hand hygiene practices in different settings such as classrooms, dormitories, and after using shared facilities. It also explored the use of hand sanitizers as an alternative when soap and water were not available. All responses were collected electronically using KoboToolbox to minimize data entry errors and ensure data quality and confidentiality (**Supplementary file 1**).

### Validity of the questionnaire

To maximize validity, the questionnaire was pre-tested on 20 respondents before distribution as a pilot study. The aim was to examine how the participants understood and responded to the questions. After the pre-test, adjustments in phrasings were made to make the questions simple to understand.

### Ethics statement

Ethical approval was granted by the KCMC University Research Ethics Review Committee under approval number Reg.2857. Participation was voluntary, with informed consent obtained from all participants. Confidentiality was maintained by using unique codes instead of names, and data were stored securely with restricted access. Participants were informed of their right to withdraw at any time without penalty.

## Statistical analysis

Data were analyzed using Statistical Package for Social Sciences (SPSS) version 27.0 software (Chicago, USA). Descriptive statistics were summarized using frequencies and percentages for categorical variables, while means and Standard Deviations (SD) were used for continuous variables. The chi-square test was employed to examine the association between categorical variables. Logistic regression analysis was conducted to determine the association between independent predictors and the outcomes of interest, such as knowledge, attitude, and practice. A *p*-value of less than 0.05 was considered significant. Knowledge was evaluated using a dichotomous scoring system, where participants received 1 point for each correct response and 0 for incorrect responses. Scores were categorized as “good knowledge” and “poor knowledge” based on a 9-item knowledge questionnaire, where participants scoring 7–9 correct responses were classified as having good knowledge, while those scoring ≤6 were classified as having poor knowledge. Attitudes were assessed using a 10-item scale, with responses measured on a Likert-type scale (1–3 ranging from “disagree” to “agree”), and overall scores were categorized as “positive attitude” (scores of 6–10) and “negative attitude” (scores ≤5). Practice was measured using a 9-item questionnaire, where correct responses were assigned 1 point and incorrect responses 0; total scores of 5–9 were categorized as “good practice,” while scores ≤4 indicated “poor practice[21].

## Results

### Demographic Characteristics of the Participants

A total of 380 students were enrolled in the study. The mean age of the participants was 23.5 ± 3.5 years. More than half of the participants 209 (55.0%) were aged 18–23 years. Females constituted a slightly higher proportion of the study population 203 (53.4%) compared to males Regarding academic programmes, BSc Laboratory Sciences (BSCL) students accounted for the largest proportion of participants 155 (40.8%), followed by Doctor of Medicine (MD) students 100 (26.3%). In terms of year of study, 115 (30.2%) were in Year 1. Participants were nearly evenly distributed according to residence, with 132 (34.9%) residing in other accommodations, 131 (34.4%) living at home, and 117 (30.7%) residing in hostels. Most participants reported having access to hand hygiene facilities 338 (88.9%). The majority had undergone clinical placement 322 (84.7%). Furthermore, nearly all participants had received formal training on hand hygiene 360 (94.7%) (Table 1).

**Table 1.** Demographic Characteristics of the Study Participants (N = 380)

| **Variables** | **Frequency (n)** | **Percentage (%)** |
| --- | --- | --- |
| **Age (years)** |  |  |
| 18–23 | 209 | 55.0 |
| 24–29 | 159 | 41.8 |
| >30 | 12 | 3.2 |
| Mean (SD) | 23.5 (3.5) |  |
| **Sex** |  |  |
| Male | 177 | 46.6 |
| Female | 203 | 53.4 |
| **Programme of Study** |  |  |
| Doctor of Medicine (MD) | 100 | 26.3 |
| Nursing | 90 | 23.7 |
| BSc Laboratory Sciences (BSCL) | 155 | 40.8 |
| Other Health Programmes | 35 | 9.2 |
| **Year of Study** |  |  |
| Year 1 | 115 | 30.2 |
| Year 2 | 98 | 25.9 |
| Year 3 | 89 | 23.3 |
| Year 4 and 5 | 78 | 20.6 |
| **Residence** |  |  |
| Home | 131 | 34.4 |
| Hostel | 117 | 30.7 |
| Other | 132 | 34.9 |
| **Access to Hand Hygiene Facilities** |  |  |
| Yes | 338 | 88.9 |
| No | 42 | 11.1 |
| **Clinical Placement** |  |  |
| Yes | 322 | 84.7 |
| No | 58 | 15.3 |
| **Formal Training on Hand Hygiene** |  |  |
| Yes | 360 | 94.7 |
| No | 20 | 5.3 |

### Knowledge regarding hand hygiene

Generally, 338 (88.9%) of participants had good knowledge regarding hand hygiene (Fig 1).

**Fig 1.**
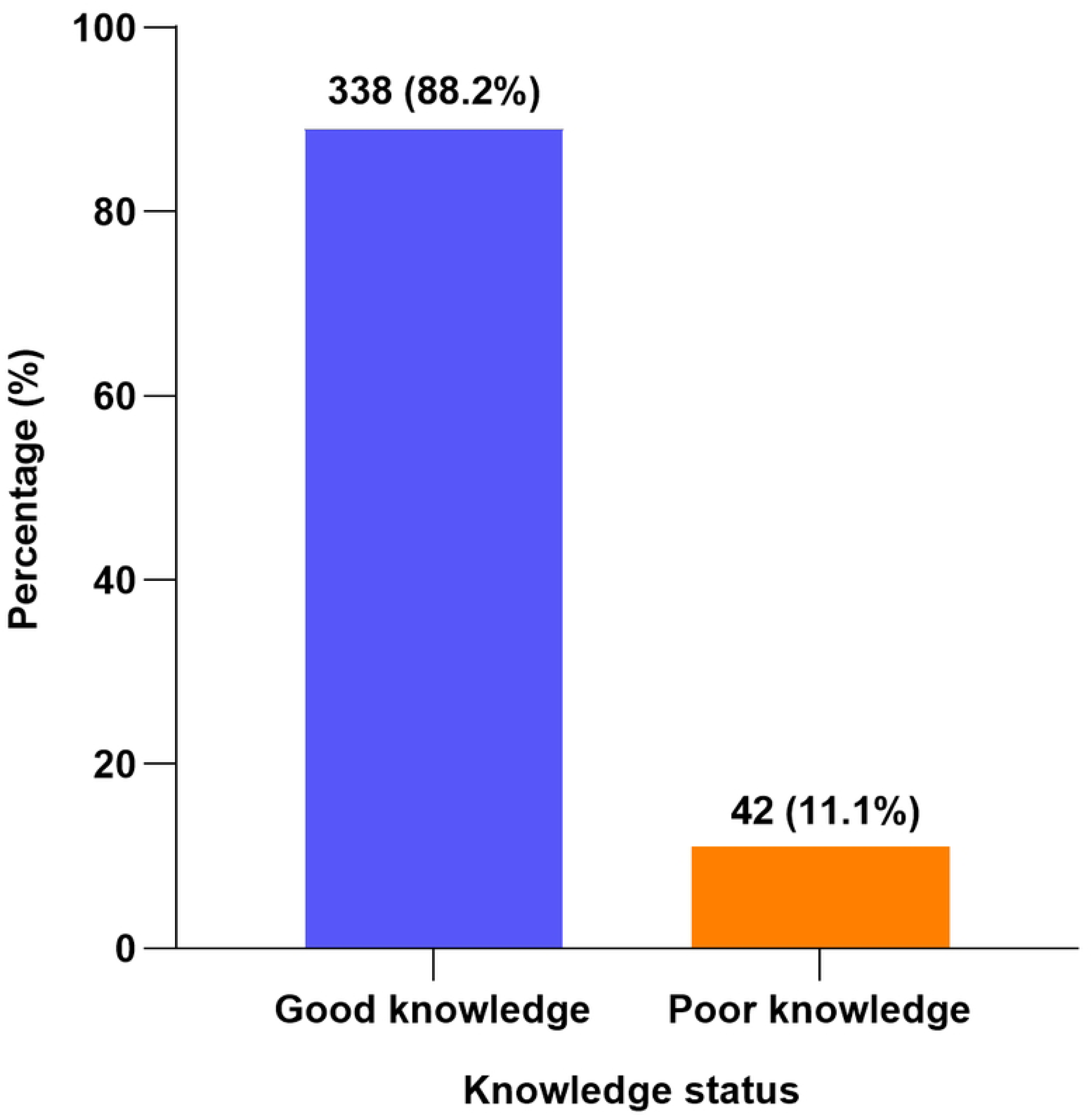
Knowledge of hand hygiene practices among university students at KCMC University, Moshi, Tanzania (N = 380).

For individual knowledge items, 134 (71.0%) of participants either strongly agreed or agreed that they knew the correct type of water recommended for proper handwashing, while 24 (12.7%) disagreed or strongly disagreed. A large proportion, 171 (90.5%), agreed that washing hands with soap and water is more effective than using water alone. Regarding hygiene practices, 165 (87.3%) agreed that watches and bracelets should be removed during handwashing, while 164 (86.7%) recognized that proper handwashing includes washing the wrists. In addition, 169 (89.4%) agreed that drying hands after washing is an essential part of hand hygiene. Most participants 175 (92.6%) reported knowing the essential materials for effective handwashing, including soap, antiseptics, and alcohol-based hand rubs. Furthermore, 169 (89.4%) agreed that proper hand hygiene can prevent infectious diseases such as COVID- 19, typhoid, and diarrhea. Regarding WHO recommendations, 153 (80.9%) of participants indicated awareness of the “5 Moments for Hand Hygiene.” In addition, 169 (89.4%) were aware of the consequences of poor hand hygiene, including its role in antimicrobial resistance (Table 2).

**Table 2.** Knowledge of Hand Hygiene Practices Among Participants (N = 380)

| S/N | Question | Strongly Agree n (%) | Agree n (%) | Neutral n (%) | Disagree n (%) | Strongly Disagree n (%) |
| --- | --- | --- | --- | --- | --- | --- |
| 1 | I know the correct type of water recommended for proper hand washing. | 149 (39.8) | 125 (32.8) | 58 (15.3) | 30 (7.9) | 18 (4.8) |
| 2 | Washing hands with soap and water is more effective than water alone. | 197 (51.9) | 147 (38.6) | 34 (9.0) | 2 (0.5) | 0 (0.0) |
| 3 | Watches and bracelets should be removed during hand washing. | 185 (48.7) | 147 (38.6) | 42 (11.1) | 6 (1.6) | 0 (0.0) |
| 4 | Proper hand washing includes washing the wrists. | 175 (46.0) | 155 (40.7) | 40 (10.6) | 4 (1.1) | 6 (1.6) |
| 5 | Drying hands after washing is essential for proper hand hygiene. | 185 (48.7) | 155 (40.7) | 34 (9.0) | 6 (1.6) | 0 (0.0) |
| 6 | I can identify materials for effective hand washing (soap, antiseptics, alcohol-based rubs). | 193 (50.8) | 159 (41.8) | 26 (6.8) | 4 (1.1) | 0 (0.0) |
| 7 | Proper hand hygiene can prevent infectious diseases such as COVID-19, typhoid, and diarrhea. | 201 (52.9) | 139 (36.5) | 38 (10.1) | 2 (0.5) | 0 (0.0) |
| 8 | I can identify the WHO 5 Moments for Hand Hygiene. | 161 (42.3) | 147 (38.6) | 52 (13.8) | 12 (3.2) | 8 (2.1) |
| 9 | I am aware of the consequences of poor hand hygiene including antimicrobial resistance. | 179 (47.1) | 160 (42.3) | 38 (10.1) | 2 (0.5) | 0 (0.0) |

## Attitude regarding hand hygiene

Generally, 193 (50.8%) of the participants expressed a negative attitude towards hand hygiene, while 187 (49.2%) demonstrated a positive attitude (Fig 2).

**Fig 2.**
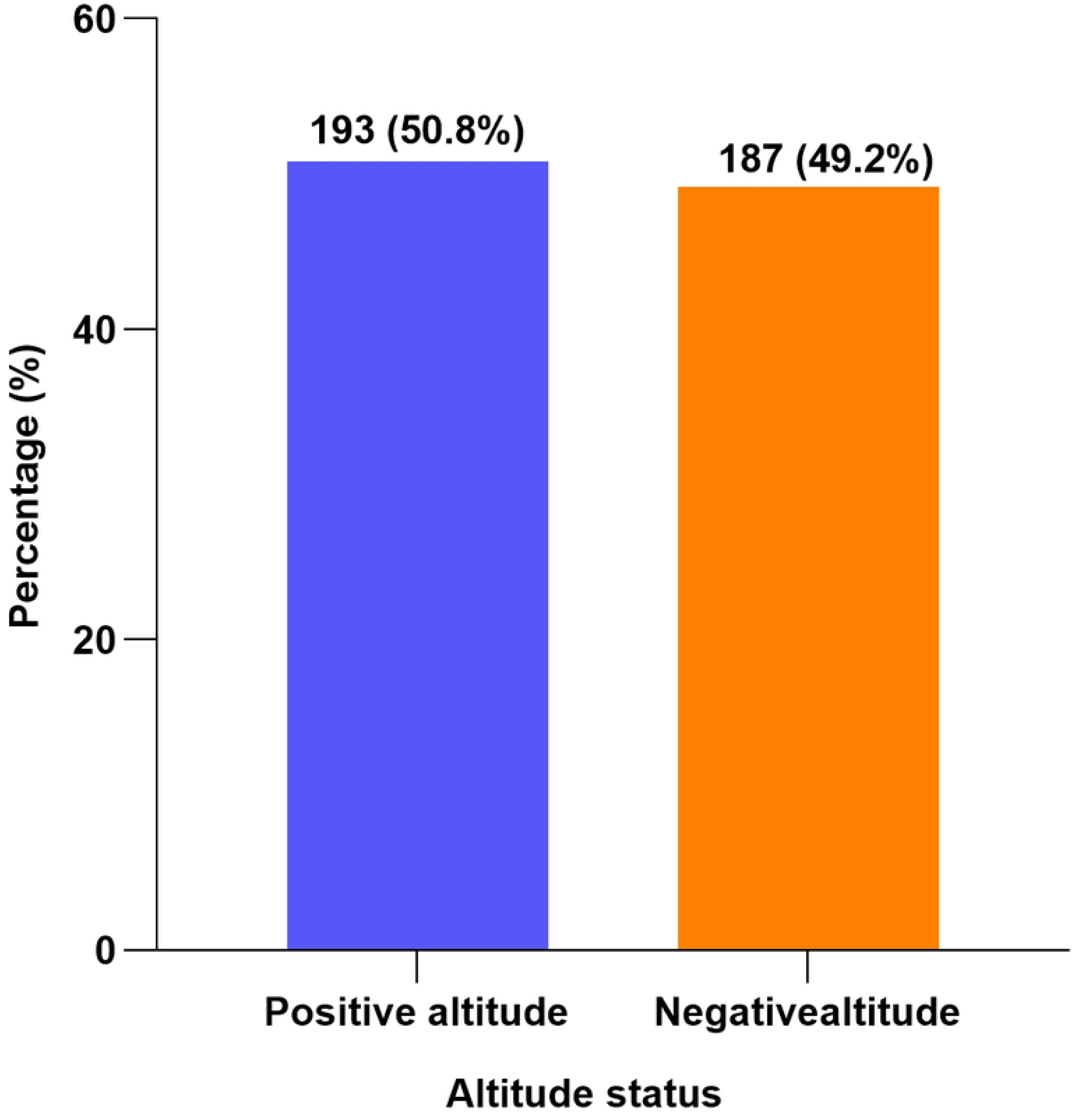
Attitudes toward hand hygiene practices among university students at KCMC University, Moshi, Tanzania (N=380).

Regarding perceptions of hand hygiene necessity, 126 (33.3%) of participants agreed or strongly agreed that handwashing after using the toilet is unnecessary if hands appear clean, while 161 (42.4%) disagreed or strongly disagreed. Similarly, 161 (42.4%) of participants agreed or strongly agreed that it is acceptable to skip handwashing when in a hurry, whereas 115 (30.2%) disagreed or strongly disagreed. Environmental and situational factors were also reported to influence hand hygiene behaviour. About 224 (58.8%) of participants indicated that availability of water significantly affects their willingness to practice hand hygiene. In addition, 193 (50.8%) agreed that handwashing is less critical when soap is not available, and 179 (47.1%) reported that they are less likely to wash hands when water feels cold or uncomfortable.

Furthermore, 153 (40.2%) of respondents indicated that handwashing after using the toilet may become irrelevant if it is not part of their routine, while 159 (41.8%) disagreed with this statement. A proportion of 154 (40.8%) agreed that insufficient drying materials may justify skipping handwashing, whereas 143 (37.6%) disagreed. On a positive note, a majority of participants demonstrated favorable attitudes towards hand hygiene promotion. Approximately 278 (73.0%) reported confidence in consistently following proper hand hygiene practices. Additionally, 338 (88.9%) believed that educating others about hand hygiene is vital for public health, and 324 (85.2%) acknowledged that attitudes and cultural norms strongly influence the adoption of hand hygiene practices (Table 3).

**Table 3.** Levels of Attitudes of Participants Towards Hand Hygiene Practices (N = 380)

| S/N |  | Strongly Agree n (%) | Agree n (%) | Neutral n (%) | Disagree n (%) | Strongly Disagree n (%) |
| --- | --- | --- | --- | --- | --- | --- |
| 1 | Hand washing after using the toilet is unnecessary if hands appear clean. | 42 (11.1) | 84 (22.2) | 92 (24.3) | 129 (33.9) | 33 (8.5) |
| 2 | It is acceptable to skip hand washing if in a hurry. | 81 (21.2) | 81 (21.2) | 105 (27.5) | 87 (22.8) | 26 (7.4) |
| 3 | Availability of water significantly impacts willingness to practice hand hygiene. | 101 (26.5) | 123 (32.3) | 72 (19.0) | 67 (17.5) | 18 (4.8) |
| 4 | Hand washing is less critical when soap is not available. | 68 (18.0) | 125 (32.8) | 84 (22.2) | 76 (20.1) | 27 (6.9) |
| 5 | I am less likely to wash hands if water feels cold or uncomfortable. | 52 (13.8) | 127 (33.3) | 95 (24.9) | 60 (15.9) | 46 (12.2) |
| 6 | Hand washing after using the toilet becomes irrelevant if not part of my routine. | 50 (13.2) | 103 (27.0) | 83 (21.7) | 81 (21.2) | 64 (16.9) |
| 7 | Insufficient drying materials justify skipping hand washing. | 48 (12.7) | 106 (28.0) | 83 (21.7) | 90 (23.8) | 53 (13.8) |
| 8 | I feel confident in my ability to consistently follow proper hand hygiene. | 121 (31.7) | 157 (41.3) | 81 (21.2) | 16 (4.2) | 6 (1.6) |
| 9 | I believe educating others about hand hygiene is vital for public health. | 159 (41.8) | 179 (47.1) | 36 (9.5) | 0 (0.0) | 6 (1.6) |
| 10. | Attitudes and cultural norms strongly influence hand hygiene practices. | 147 (38.6) | 177 (46.6) | 48 (12.7) | 6 (1.6) | 2 (0.5) |

## Practices of Participants Towards Hand Hygiene

Generally, 201 (52.9%) of the participants engaged in poor hand hygiene practices, while 179 (47.1%) demonstrated good practices (Fig 3).

**Fig 3.**
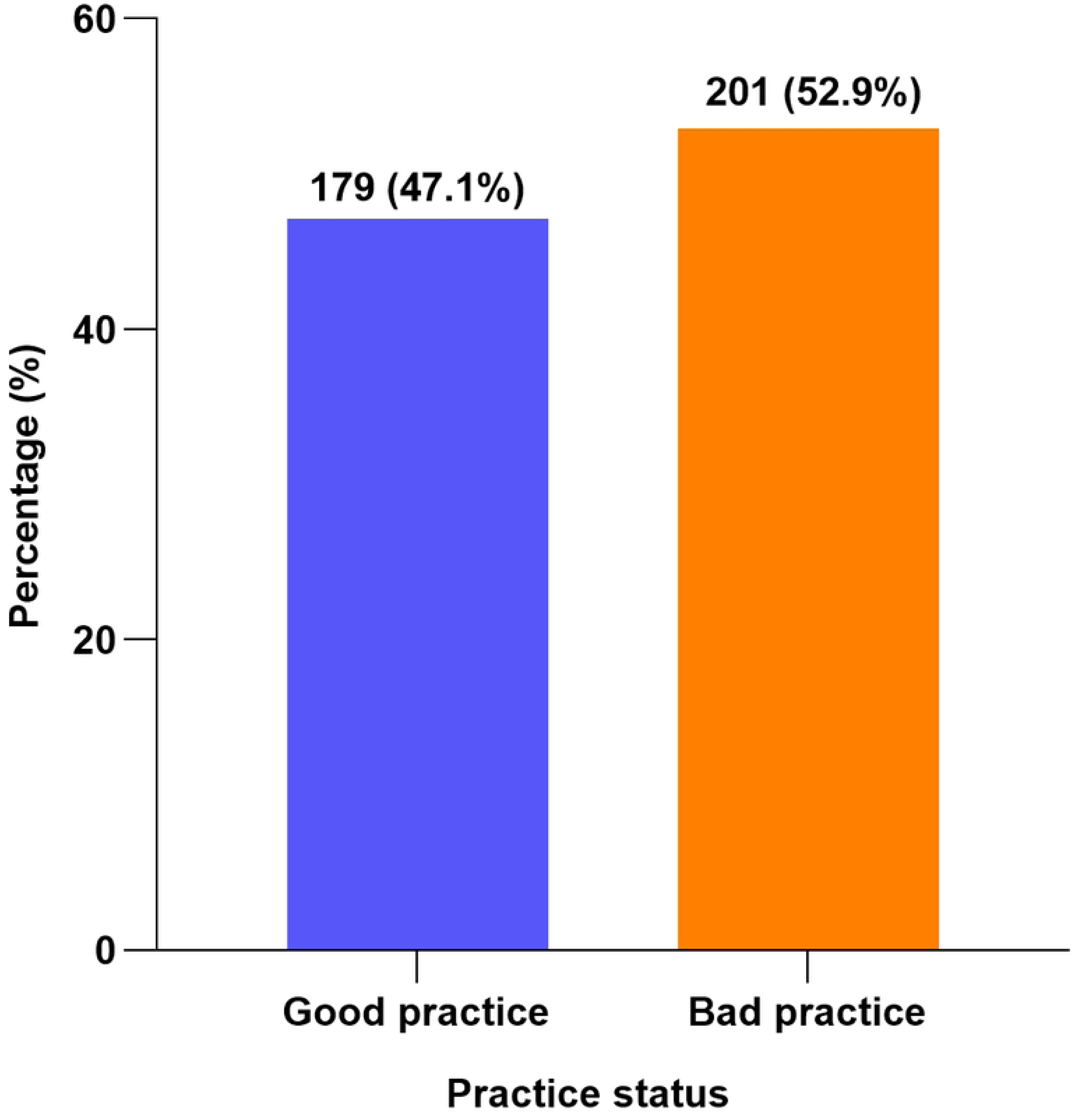
Hand hygiene practices among university students at KCMC University, Moshi, Tanzania (N = 380).

Regarding general hand hygiene practices, 131 (34.4%) of participants reported regular use of sanitizers as part of their routine, while 147 (38.6%) reported not using sanitizers regularly. Only 67 (17.5%) reported washing hands after visiting public places such as markets, churches, or mosques, whereas 205 (54.0%) did not practice this consistently. Similarly, 83 (21.7%) prioritized handwashing after touching commonly shared surfaces such as ATMs or money, while 189 (49.7%) did not. With respect to home and daily life practices, 99 (25.9%) reported washing hands immediately upon returning home after being outside, while 147 (38.6%) did not. In addition, 143 (37.6%) reported following proper hand hygiene before and after eating, whereas 62 (16.4%) did not.

In clinical-related practices, a relatively higher proportion of participants demonstrated good compliance. A total of 261 (68.8%) reported washing hands thoroughly after using the toilet, and 265 (69.8%) indicated adherence to proper hand hygiene during clinical or practical sessions, including before and after wearing gloves. Furthermore, 185 (48.7%) reported performing hand hygiene immediately after exposure to body fluids during patient care, while 74 (19.6%) did not. Additionally, 318 (83.6%) reported washing hands after contact with patient surroundings such as bed rails or medical equipment (Table 4).

**Table 4.** Levels of Hand Hygiene Practices Among Participants (N = 380)

| S/N | Question | Agree n (%) | Disagree n (%) | Not Sure n (%) |
| --- | --- | --- | --- | --- |
| 1 | Use of sanitizers regularly as part of hand hygiene routine | 131 (34.4) | 147 (38.6) | 102 (27.0) |
| 2 | Washing hands after visiting public places (markets, churches, mosques) | 67 (17.5) | 205 (54.0) | 108 (28.6) |
| 3 | Prioritizing handwashing after touching surfaces like ATMs or money | 83 (21.7) | 189 (49.7) | 108 (28.6) |
| 4 | Washing hands immediately after returning home from outside | 99 (25.9) | 147 (38.6) | 134 (35.4) |
| 5 | Following proper hand hygiene before and after eating | 143 (37.6) | 62 (16.4) | 175 (46.0) |
| 6 | Washing hands thoroughly after using toilets | 261 (68.8) | 60 (15.9) | 58 (15.3) |
| 7 | Ensuring hand hygiene during clinical/practical sessions (before and after gloves use) | 265 (69.8) | 64 (16.9) | 50 (13.2) |
| 8 | Performing hand hygiene after exposure to body fluids during patient care | 185 (48.7) | 74 (19.6) | 121 (31.7) |
| 9 | Washing hands after contact with patient surroundings (bed rails, equipment) | 318 (83.6) | 44 (11.6) | 18 (4.8) |

## Factors associated with knowledge, attitudes and practice regarding hand hygiene

Multivariable logistic regression analysis identified several factors significantly associated with hand hygiene knowledge, attitudes, and practices. Good knowledge was independently associated with being in the fourth year or above (AOR = 2.31, 95% CI: 1.07–4.99, *p* = 0.033), access to hand hygiene facilities (AOR = 2.84, 95% CI: 1.25–6.42, *p* = 0.012), clinical placement experience (AOR = 2.21, 95% CI: 1.01–4.85, *p* = 0.047), formal training on hand hygiene (AOR = 3.48, 95% CI: 1.24–9.76, *p* = 0.018), and enrolment in the BSc Laboratory Sciences programme (AOR = 2.18, 95% CI: 1.15–4.13, *p* = 0.017) (Table 5). Positive attitudes towards hand hygiene were significantly associated with being in the fourth year or above (AOR = 2.12, 95% CI: 1.16–3.89, *p* = 0.015), access to hand hygiene facilities (AOR = 2.16, 95% CI: 1.04–4.50, *p* = 0.040), and clinical placement experience (AOR = 1.89, 95% CI: 1.01–3.55, *p* = 0.048) (Table 6).

**Table 5:**
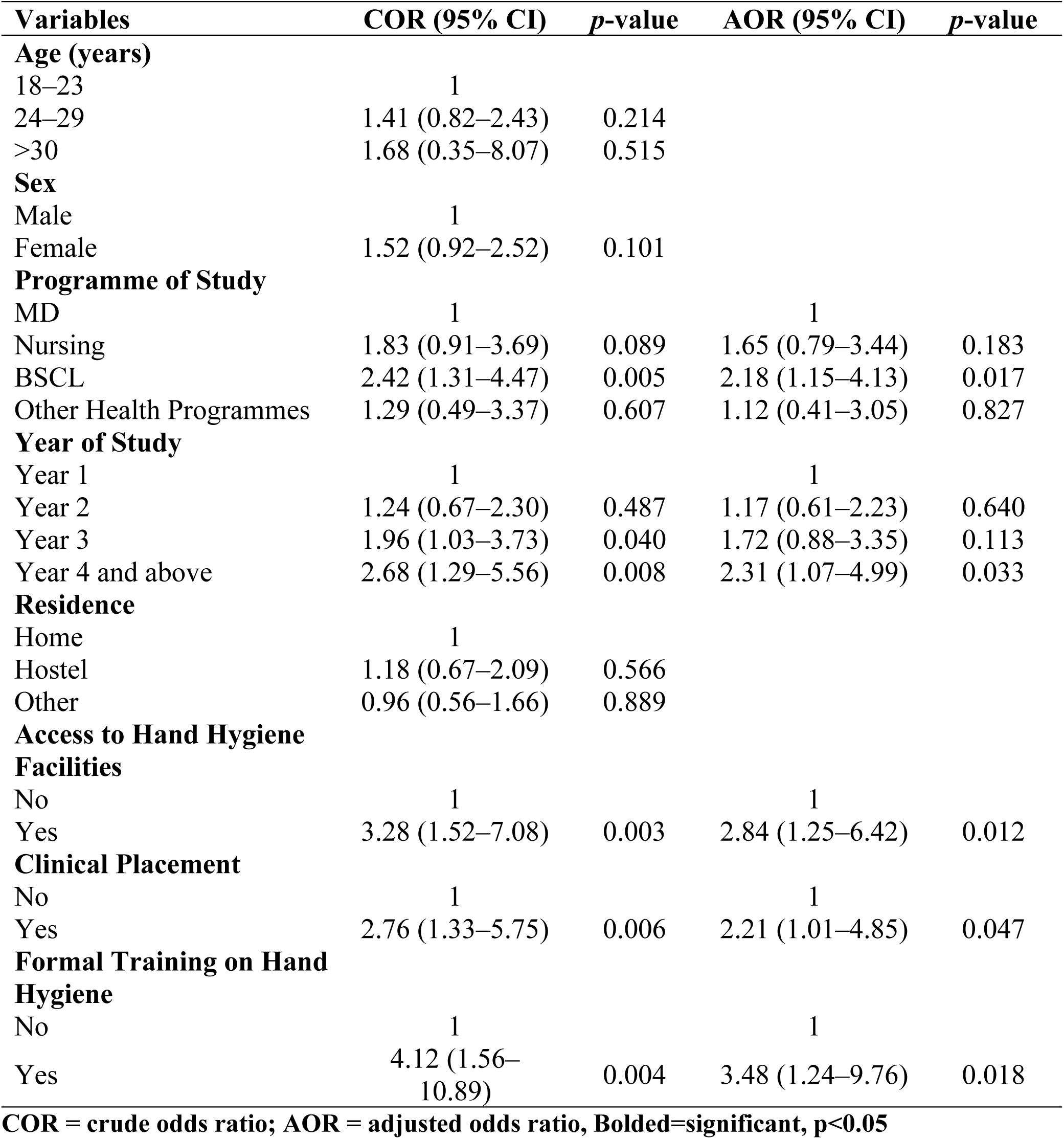
Factors associated with knowledge regarding hand hygiene (N=380)

**Table 6:** Factors associated with attitudes regarding hand hygiene (N=380)

| **Variables** | **COR (95% CI)** | ***p*-value** | **AOR (95% CI)** | ***p*-value** |
| --- | --- | --- | --- | --- |
| **Age (years)** |  |  |  |  |
| 18–23 | 1 |  |  |  |
| 24–29 | 1.36 (0.95–1.96) | 0.095 |  |  |
| >30 | 1.74 (0.61–4.99) | 0.301 |  |  |
| **Sex** |  |  |  |  |
| Male | 1 |  | 1 |  |
| Female | 1.49 (1.01–2.19) | 0.043 | 1.34 (0.89–2.03) | 0.160 |
| **Programme of Study** |  |  |  |  |
| MD | 1 |  | 1 |  |
| Nursing | 1.69 (1.01–2.84) | 0.046 | 1.51 (0.87–2.62) | 0.144 |
| BSCL | 1.84 (1.14–2.95) | 0.012 | 1.63 (0.98–2.72) | 0.058 |
| Other Health Programmes | 1.05 (0.49–2.25) | 0.905 | 0.97 (0.43–2.17) | 0.936 |
| **Year of Study** |  |  |  |  |
| Year 1 | 1 |  | 1 |  |
| Year 2 | 1.26 (0.78–2.05) | 0.349 | 1.18 (0.70–1.99) | 0.532 |
| Year 3 | 1.81 (1.08–3.03) | 0.024 | 1.67 (0.97–2.87) | 0.064 |
| Year 4 and above | 2.45 (1.39–4.33) | 0.002 | 2.12 (1.16–3.89) | 0.015 |
| **Residence** |  |  |  |  |
| Home | 1 |  |  |  |
| Hostel | 1.11 (0.72–1.72) | 0.630 |  |  |
| Other | 0.92 (0.60–1.42) | 0.712 |  |  |
| **Access to Hand Hygiene Facilities** |  |  |  |  |
| No | 1 |  | 1 |  |
| Yes | 2.57 (1.30–5.06) | 0.006 | 2.16 (1.04–4.50) | 0.040 |
| **Clinical Placement** |  |  |  |  |
| No | 1 |  | 1 |  |
| Yes | 2.12 (1.17–3.84) | 0.013 | 1.89 (1.01–3.55) | 0.048 |
| **Formal Training on Hand Hygiene** |  |  |  |  |
| No | 1 |  | 1 |  |
| Yes | 2.68 (1.02–7.05) | 0.046 | 2.11 (0.77–5.77) | 0.146 |

**COR = crude odds ratio; AOR = adjusted odds ratio, Bolded=significant, $p < 0.05$**

Similarly, good hand hygiene practices were independently associated with female sex (AOR = 1.58, 95% CI: 1.01–2.48, *p* = 0.045), enrolment in the BSc Laboratory Sciences programme (AOR = 1.97, 95% CI: 1.10–3.52, *p* = 0.022), being in the third year (AOR = 1.89, 95% CI: 1.02–3.49, *p* = 0.042) or fourth year and above (AOR = 2.41, 95% CI: 1.19–4.88, *p* = 0.014), access to hand hygiene facilities (AOR = 2.67, 95% CI: 1.18–6.01, *p* = 0.018), clinical placement experience (AOR = 2.05, 95% CI: 1.03–4.08, *p* = 0.040), and formal training on hand hygiene (AOR = 2.98, 95% CI: 1.19–7.48, *p* = 0.020) (Table 7).

**Table 7:** Factors associated with practice regarding hand hygiene (N=380)

| **Variables** | **COR (95% CI)** | **p-value** | **AOR (95% CI)** | **p-value** |
| --- | --- | --- | --- | --- |
| **Age (years)** |  |  |  |  |
| 18–23 | 1 |  |  |  |
| 24–29 | 1.32 (0.89–1.97) | 0.167 |  |  |
| >30 | 1.58 (0.52–4.80) | 0.420 |  |  |
| **Sex** |  |  |  |  |
| Male | 1 |  | 1 |  |
| Female | 1.72 (1.12–2.65) | 0.013 | 1.58 (1.01–2.48) | 0.045 |
| **Programme of Study** |  |  |  |  |
| MD | 1 |  | 1 |  |
| Nursing | 1.45 (0.82–2.56) | 0.201 | 1.21 (0.65–2.24) | 0.549 |
| BSCL | 2.18 (1.28–3.69) | 0.004 | 1.97 (1.10–3.52) | 0.022 |
| Other Health Programmes | 1.12 (0.48–2.61) | 0.792 | 0.98 (0.40–2.41) | 0.963 |
| **Year of Study** |  |  |  |  |
| Year 1 | 1 |  | 1 |  |
| Year 2 | 1.34 (0.76–2.36) | 0.312 | 1.25 (0.69–2.26) | 0.456 |
| Year 3 | 2.08 (1.16–3.74) | 0.014 | 1.89 (1.02–3.49) | 0.042 |
| Year 4 and above | 2.84 (1.49–5.42) | 0.002 | 2.41 (1.19–4.88) | 0.014 |
| **Residence** |  |  |  |  |
| Home | 1 |  |  |  |
| Hostel | 1.17 (0.73–1.87) | 0.516 |  |  |
| Other | 0.94 (0.59–1.51) | 0.806 |  |  |
| **Access to Hand Hygiene Facilities** |  |  |  |  |
| No | 1 |  | 1 |  |
| Yes | 2.94 (1.36–6.37) | 0.006 | 2.67 (1.18–6.01) | 0.018 |
| **Clinical Placement** |  |  |  |  |
| No | 1 |  | 1 |  |
| Yes | 2.31 (1.21–4.41) | 0.011 | 2.05 (1.03–4.08) | 0.040 |
| **Formal Training on Hand Hygiene** |  |  |  |  |
| No | 1 |  | 1 |  |
| Yes | 3.45 (1.44–8.26) | 0.005 | 2.98 (1.19–7.48) | 0.020 |

**COR = crude odds ratio; AOR = adjusted odds ratio, Bolded=significant, p<0.05**

## Discussion

The present study revealed that although university students demonstrated high knowledge of hand hygiene (88.9%), this did not consistently translate into positive attitudes and good practices, as 50.8% reported negative attitudes and 52.9% exhibited poor practices. This persistent knowledge–practice gap has been widely reported in studies among university students and healthcare trainees, indicating that knowledge alone is insufficient to ensure behavioural change [22,23].

The high level of knowledge observed in this study is consistent with findings from East Africa, where generally good awareness of hand hygiene has been reported among students and healthcare workers. A study conducted in Tanzania reported high awareness of infection prevention measures among university students in Mwanza during the COVID-19 pandemic [23]. Similarly, a study conducted in Uganda found that more than three-quarters of medical students demonstrated adequate knowledge of hand hygiene principles[20]. These findings suggest that while awareness levels are relatively high across the region, this does not necessarily translate into consistent hand hygiene practices.

Despite high knowledge, the current study revealed poor hand hygiene practices (52.9%), which is consistent with findings from Tanzania and other low- and middle-income countries. In Uganda, compliance with hand hygiene among university students remained suboptimal even during the COVID-19 pandemic, indicating that increased awareness did not necessarily translate into sustained behavioural change [24]. Similarly, a study conducted in Bangladesh among university students reported that although knowledge of hand hygiene was high, less than half of participants adhered to recommended handwashing practices due to inadequate facilities and behavioural factors [15]. Likewise, a study in India found a significant gap between knowledge and practice of hand hygiene among undergraduate students, with infrastructural limitations and low risk perception identified as key barriers [16].

The decline in proper hand hygiene practice in the post-COVID-19 era has also been reported in recent studies, suggesting behavioural fatigue following public health emergencies[9] .A study done by Lutainulwa et al. (2026) observed reduced adherence to infection prevention and control practices among healthcare workers in Dar es Salaam after the COVID-19 peak period, indicating that improvements achieved during outbreaks are often not sustained over time without continuous reinforcement[25]. This highlights the need for ongoing training and reinforcement strategies to maintain adherence to infection prevention measures beyond emergency periods.

The disconnect between knowledge and practice observed in this study is also supported by international evidence highlighting that behavioural, institutional, and environmental factors strongly influence compliance [26]. A study conducted in South Africa reported that although students had good knowledge and positive attitudes toward hand hygiene, their practices remained inconsistent due to structural and behavioural barriers [22]. Similarly, a study conducted in Kenya identified lack of adequate water, soap, and sanitizers as major constraints to proper hygiene practice among students despite adequate knowledge [27,28]. These findings emphasise that environmental and infrastructural limitations play a critical role in determining hand hygiene compliance beyond knowledge alone.

In the present study, significant factors associated with knowledge, attitudes, and practices included access to hand hygiene facilities, formal training, clinical placement, academic year, and programme of study. Access to hygiene facilities has consistently been identified as a key determinant of compliance in previous studies [23,26].Similarly, students in higher academic years and those with clinical exposure tend to demonstrate better hand hygiene behaviour due to repeated reinforcement and practical experience[15,20,29]

Formal training on infection prevention was also strongly associated with better knowledge and practice in this study, consistent with findings from previous research [11,16]. Training enhances both awareness and behavioural reinforcement, leading to improved compliance with recommended hand hygiene practices [10,11,30]. Additionally, the programme of study influenced hand hygiene behaviour, with health-related disciplines such as laboratory sciences demonstrating better practices compared to other programmes, consistent with findings from similar academic settings [22].

Generally, the findings of this study are consistent with previous literature showing a persistent gap between knowledge and practice of hand hygiene among university students. Despite high levels of knowledge, actual practices remain suboptimal and are largely influenced by access to facilities, training, and clinical exposure. This study highlights that knowledge alone is insufficient to sustain proper hand hygiene behaviour. These results underscore the need for integrated interventions combining health education, adequate infrastructure, and continuous behavioural reinforcement to improve compliance in university settings.

## Strengths and limitations

This study provides important and timely evidence on hand hygiene knowledge, attitudes, and practices among university students in the post-COVID-19 era, a period where sustaining infection prevention behaviors remains a public health concern. The study benefits from the use of both descriptive and inferential statistical approaches, including logistic regression and multivariable analysis, which improve the robustness of identifying factors associated with hand hygiene practices. Focusing on university students in a high-contact academic environment enhances the relevance of the findings for targeted interventions and infection prevention and control programs in similar settings. However, the study is limited by its reliance on self-reported data, which may be affected by recall bias and social desirability bias, potentially leading to overestimation of good practices. In addition, the cross-sectional design does not allow for determination of causal relationships between knowledge, attitudes, and practices. Finally, being conducted in a single university limits the generalizability of the findings to other universities or broader populations in different geographical and socio- economic contexts.

## Conclusion

This study demonstrates that although university students at KCMC University have generally high knowledge of hand hygiene and recognize its importance in preventing infectious diseases, this knowledge is not consistently reflected in their attitudes and practices. A substantial proportion of students still exhibit negative attitudes and poor hand hygiene practices in the post-COVID-19 era, highlighting a persistent gap between knowledge and behaviour. This gap is consistent with findings from other studies, which show that knowledge alone is insufficient to ensure adherence to proper hand hygiene practices. The observed outcomes further indicate that factors such as access to hand hygiene facilities, formal training, and clinical exposure play an important role in influencing better knowledge, attitudes, and practices. Therefore, sustained and targeted health promotion strategies, improved access to hygiene facilities, and continuous behavioural reinforcement are necessary to bridge this gap and reduce the risk of infectious disease transmission among university students.

## Data Availability

All data can be found in the manuscript and supporting information files.

## List of Abbreviations

AOR: Adjusted Odds Ratio
CDC: Centers for Disease Control and Prevention
CI: Confidence Interval
COVID-19: Coronavirus Disease 2019
COR: Crude Odds Ratio
HAIs: Healthcare-Associated Infections
KAP: Knowledge, Attitudes and Practices
LMIC: Low- and Middle-Income Countries
SARS-CoV-2: Severe Acute Respiratory Syndrome Coronavirus 2
UNICEF: United Nations Children’s Fund
WHO: World Health Organization

## Acknowledgements

We express our gratitude to the study participants for their willingness to participate.

## Author Contributions

**Conceptualization:** Emmanuel T. Issangya, Debora C. Kajeguka, Rhodes Mwageni **Methodology:** Emmanuel T. Issangya, Debora C. Kajeguka, Rhodes Mwageni **Data curation:** Emmanuel T. Issangya, Dhahiri Mnzava

**Formal analysis:** Emmanuel T. Issangya, Dhahiri Mnzava **Investigation:** Emmanuel T. Issangya, Dhahiri Mnzava **Writing – original draft:** Emmanuel T. Issangya

**Writing – review & editing:** Emmanuel T. Issangya, Victor V. Mosha, Dhahiri Mnzava, Rhodes Mwageni, Debora C. Kajeguka

**Supervision:** Debora C. Kajeguka, Rhodes Mwageni

**Project administration:** Emmanuel T. Issangya, Debora C. Kajeguka, Rhodes Mwageni

## Notes

### Competing Interest Statement

The authors have declared no competing interest.

### Clinical Trial

NA

### Author Declarations

Ethical approval was granted by the KCMC University Research Ethics Review Committee under approval number Reg.2857

